# Settings, Characteristics, and Experiences of Stigma Among People with Tuberculosis in Kenya: National Survey Results

**DOI:** 10.1101/2025.09.22.25336407

**Authors:** Aiban Ronoh, Joshua Limo, Lazarus Odeny, Stephen Macharia, Rachael Muinde, Joyce Kiarie, Jane Ong’ang’o, Nkirote Mwirigi, Drusilla Nyaboke, Evaline Kibuchi, Timothy Kilonzo, Rose Wandia, L. Nkirote Mugambi-Nyaboga, Immaculate Kathure

## Abstract

**Background:** Tuberculosis (TB)-related stigma remains a significant barrier to TB care and treatment adherence in Kenya. Despite progress in TB control, stigma continues to affect individuals diagnosed with TB and their families, leading to delayed healthcare-seeking behaviors, social exclusion, and economic consequences. This study examines the dimensions of TB-related stigma among people infected and affected by TB in Kenya.

**Methods:** A cross-sectional study was conducted in November 2023. Data were collected from people with TB (PWTB) in 180 health facilities, across 11 counties in Kenya. A multistage stratified sampling method was employed to ensure regional representation. Stigma levels were assessed using a structured stigma index, and multivariate logistic regression was used to analyze factors associated with stigma.

**Results:** A total of 367 PWTB were included in the analysis. Most (67%) were men and the median age was 35 years. The study found high levels of community stigma (68%), family stigma (52%), healthcare system stigma (51%) and self-stigma (49%). Many PWTB reported concealing their diagnosis due to fear of discrimination, while families and communities often distanced themselves from individuals with TB.

**Conclusion:** TB-related stigma in Kenya is prevalent across multiple dimensions, affecting individuals, families, communities, and healthcare systems. Addressing stigma requires targeted interventions, including awareness campaigns, stigma reduction training for healthcare workers, and policy reforms to promote inclusive and supportive TB care environments.

## 1. Introduction

Kenya is among the 30 high TB-burden countries that contribute to 80% of the global burden of TB disease (1) WHO estimates that 124,000 people fell ill due to tuberculosis in Kenya in 2023 but only 97,126 TB cases were diagnosed and started on treatment. This represented a case detection rate of 77% with 23% of TB cases being missed (1). The overall case notification rate for the year 2023 was 204 TB cases per 100,000 population(2), well below the estimated case notification rate established during the prevalence survey conducted in 2016 (3). This is further indication that there are people with TB missed within communities and the health system.

Tuberculosis control efforts have increasingly become patient-centered, emphasizing universal access to care. A major milestone was the 2018 UN General Assembly high-level meeting on TB, where a political declaration was endorsed to accelerate progress toward ending TB by 2035. Among its key objectives was the elimination of stigma and discrimination, recognizing that addressing stigma is central to creating a supportive environment for people with TB and improving overall TB control (4).

Stigma, described as a process of devaluation, involves being discredited, viewed as shameful, or perceived as threatening (5). Globally, stigma is a well-documented barrier to health-seeking, engagement in care, and treatment adherence (6) (7)(8). Misconceptions about TB transmission and its association with HIV/AIDS fuel social isolation and discrimination. Families may isolate members with TB, particularly women, due to fear of infection and social consequences (Mbuthia et al., 2020). Negative attitudes from healthcare providers and operational barriers within health systems further exacerbate stigma, discouraging timely care-seeking (9).

Community settings are widely documented as primary loci of TB stigma, where fear of infection and misinformation drive gossip, social isolation, and rejection (10) (11). In Kenya, delayed TB care has been linked to stigma and gender norms, particularly among women and rural dwellers, who face heightened discrimination within families and communities (12) (13) (14). Similarly, in Ghana, stigma has been shown to contribute to concealment of symptoms and delays in diagnosis, particularly among women (15).

Socioeconomic factors such as low education, poverty, and lack of awareness consistently emerge as key determinants of TB stigma. Patients with low or no income report higher internalized stigma, including shame and exclusion, which undermines treatment adherence (16) (17). However, evidence is mixed: a Ugandan study of over 33,000 participants found higher stigma levels among older adults, urban residents, those with higher education, and those with greater TB knowledge, suggesting that awareness alone may not mitigate stigma and can sometimes reinforce it through heightened associations (18). Religious beliefs and faith-based organizations (FBOs), meanwhile, have demonstrated potential in reducing stigma through psychosocial support and community education (19) (20).

Stigma intensity also varies by treatment phase and drug resistance status. Newly diagnosed patients and those with drug-resistant TB (DR-TB) often report greater stigma due to prolonged treatment, visible side effects, and fear of infecting others (21) (22) (23). Anticipated stigma that is, fear of rejection if status is disclosed, has been associated with delayed diagnosis, missed appointments, and poor psychological health (22).

Within health facilities, discriminatory practices have also been reported, including negative attitudes by health workers who may blame patients for late presentation or visibly segregate them. Such actions reinforce stigma despite TB being curable (9). Understanding the origins of TB stigma is therefore integral to reducing its impact on care-seeking behaviors, which often leads to delayed diagnosis, complications, and TB-related mortality (12). Evidence from Uganda highlights high levels of TB-related stigma with notable geographic disparities, driven by community perceptions, misconceptions, and knowledge gaps (11). Similarly, in West Pokot, Kenya, stigma was strongly associated with HIV/AIDS and fear of transmission, leading to both perceived and enacted stigma (18).

Overall, TB stigma represents a powerful social label that alters self-perception and social standing. It delays care-seeking, prolongs infectiousness, worsens health outcomes, and drives self-isolation and premature treatment discontinuation (12) (24) (9). Misconceptions about the causes of TB and its transmission further exacerbate isolation and discrimination (25).

All in all, the literature demonstrates that TB stigma is multidimensional—rooted in social, economic, cultural, and institutional factors. Yet, to date, there has been no nationally representative survey of TB-related stigma in Kenya. This study, therefore sought to fill that gap by assessing the levels and dimensions of stigma experienced by people with TB, generating evidence to inform targeted interventions and contribute to the broader goal of ending TB in Kenya.

### Conceptual Framework

Figure 1 below shows the conceptual framework that was adopted during the TB stigma index survey in Kenya. It outlines the effects of stigma as perceived by affected persons within the dimensions of health seeking and social perceptions, TB prevention, TB adherence and care, and subsequent TB treatment outcomes.

**Figure 1.**
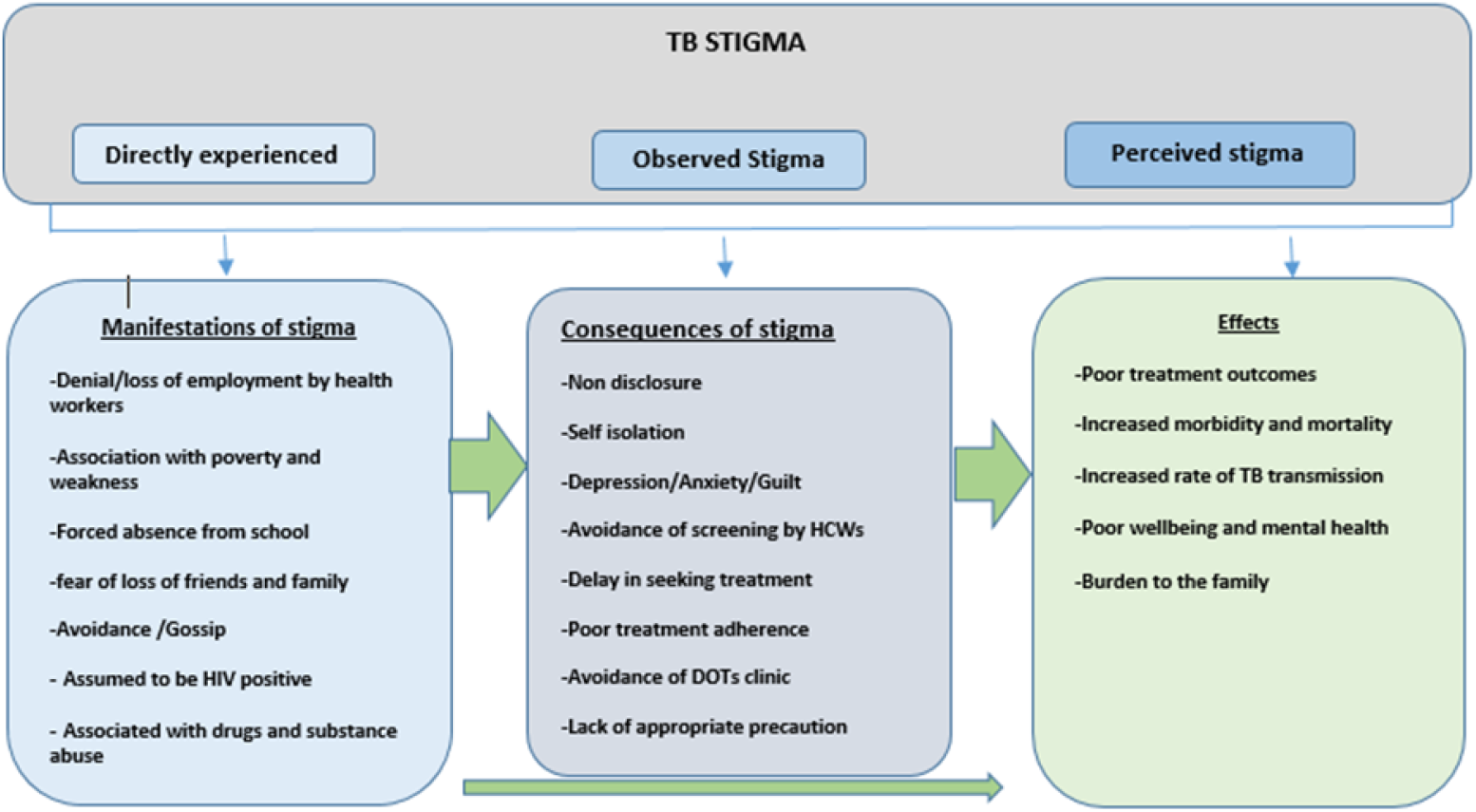
TB stigma index survey conceptual framework (Adapted from Turan & Nyblade) (20)

## 2. Methodology

### Ethics statement

Ethical approval was obtained from **Amref Health Africa Ethics and Scientific Review Committee (ESRC), approval number of being-AMREF-ESRC P1354**|**2022** and the National Commission for Science and Technology Innovation (NACOSTI|P|23|29356). The study approval covered a period of one year from 11^th^ May 2023 to 11^th^ May 2024. There was no extension sought.

### Study Design

This was a cross-sectional national survey that assessed the TB stigma index among persons with TB disease within households, communities and healthcare settings.

#### Study Setting

Kenya is administratively divided into 47 counties grouped into 12 regions for planning and coordination purposes. These counties vary in population size, TB burden, and health system capacity. While some counties, such as Nairobi, Mombasa, and Kisumu, report high TB notification rates and are considered high-burden areas, others have relatively low TB case numbers. HIV prevalence, a key driver of TB, also varies widely across counties—from under 2% in some regions to over 15% in others. Kenya’ s health system is decentralized and structured across six levels of care: community health services (Level 1), primary healthcare facilities (Levels 2–3), county referral hospitals (Level 4), and national referral hospitals (Levels 5–6). TB diagnostic services are available at multiple levels, with smear microscopy, Xpert MTB/RIF, Trunat and digital chest X-rays widely distributed. Advanced diagnostics, including TB culture and drug susceptibility testing (DST), are available at designated regional and national reference laboratories.

#### Study population

The target population comprised people with TB who were active on treatment and were 15 years and above.

#### Sampling Procedures

A multistage stratified sampling method was employed to ensure regional representation across Kenya. The country was stratified into 12 regions: Nairobi, Central, Coast, Eastern North, Eastern South, North Eastern, Rift Valley North, Rift Valley South, Nyanza North, Nyanza South, and Western. From each region, one county was purposely selected based on operational feasibility and TB burden. Within each selected county, two sub-counties were further purposely chosen. Criteria for sub-county selection included; TB case notification rates (CNR), urban–rural setting, availability of private health facilities providing TB care and presence of prison health facilities

Within these sub-counties, a total of 180 health facilities offering TB treatment services were identified as primary recruitment sites for participants undergoing TB treatment.

A sample size of 421 persons with TB was estimated using the 2021 national TB case notification figure (77,854 cases), assuming a 95% confidence level and a 90% response rate. Participants were selected consecutively from TB treatment registers at the selected facilities until the required sample size was reached.

#### Eligibility Criteria

##### Inclusion criteria

All people with tuberculosis aged 15 years and older, who gave informed consent were included in the study.

##### Exclusion criteria

All people who were below the age of 15 years were excluded and individuals under the influence of alcohol or any substance likely to impair logical reasoning at the time of interview in addition those who declined to give consent in the study.

#### Data Collection tools and procedures

Data were collected using a digital questionnaire developed on Open Data Kit (ODK). Trained data collectors administered the tool to eligible participants who provided informed consent. The completed forms were uploaded to a secure web-based system, from which data were extracted, cleaned, and exported to Excel for preliminary review.

Stigma was assessed using a set of standardized questions rated on a **five-point Likert scale** (0 = Strongly Disagree to 4 = Strongly Agree). A total stigma score was computed for each participant by summing individual item scores. These scores were further transformed into a binary variable: participants with scores less than or equal to the neutral midpoint (“ Disagree” × number of items) were categorized as having **“ no stigma**,**”** while those scoring above this threshold were categorized as experiencing **“ stigma.”**

#### Data Analysis

Exploratory factor analysis (EFA) was conducted to test the internal consistency and construct validity of the stigma scale in the Kenyan context. Cronbach’ s alpha was calculated to assess internal reliability, with values ≥0.7 indicating acceptable consistency. The principal components extraction method was used to identify underlying factors, with factor loadings ≥0.4 considered acceptable.

Data were analyzed using R statistical software (version 5.4.0) with relevant packages (e.g., xfun::pkg_attach()) for data management and reporting. A working dataset was created by subsetting and recoding key variables including: i) Sociodemographic factors: age group, sex, marital status, education, religion, occupation, place of residence, and family size and ii) psychosocial and clinical variables: perceived social support, substance use, treatment phase, patient type, mental illness, comorbidities, TB type, and stigma setting

Stigma-related responses covering domains such as guilt, fear, social avoidance, and disclosure concerns were numerically encoded (1 = Strongly Agree to 5 = Strongly Disagree). Scores were aggregated row-wise per participant to generate a mean stigma score, which was then categorized as follows: 1.00–1.49: Strongly Disagree, 1.50–2.49: Disagree, 2.50–3.49: Neutral, 3.50–4.49: Agree and 4.50–5.00: Strongly Agree.

For binary classification, participants who “ Agreed” or “ Strongly Agreed” with stigma-related items were categorized as experiencing stigma, while those who “ Disagreed” or “ Strongly Disagreed” were categorized as not experiencing stigma. Responses with a “ Neutral” score were treated as missing in the binary variable.

Descriptive statistics and cross-tabulations were generated, and inferential analysis was performed using Chi-square tests and binary logistic regression. Variables with p-values <0.05 in bivariate analysis were entered into a multivariate logistic regression model to identify independent predictors of TB-related stigma. Outputs are presented in Table 1 and Table 2 of the Results section.

**Table 1:** Demographic characteristics of people with TB.

| Characteristic | N | N = 367<sup>1</sup> |
| --- | --- | --- |
| **Age** |  |  |
| 0-14 |  | 10 (2.7%) |
| 15-24 |  | 58 (16%) |
| 25-34 |  | 110 (30%) |
| 35-44 |  | 92 (25%) |
| 45-54 |  | 51 (14%) |
| 55-64 |  | 28 (7.6%) |
| 65+ |  | 18 (4.9%) |
| **Sex** |  |  |
| Male |  | 246 (67%) |
| Female |  | 120 (33%) |
| Intersex |  | 1 (0.3%) |
| **Marital Status** |  |  |
| Single |  | 115 (31%) |
| Married |  | 182 (50%) |
| Divorced |  | 10 (2.7%) |
| Widowed |  | 21 (5.7%) |
| Separated |  | 29 (7.9%) |
| Not Applicable |  | 10 (2.7%) |
| **Education Level** |  |  |
| No formal education |  | 36 (9.8%) |
| Primary level |  | 155 (42%) |
| Secondary Level |  | 135 (37%) |
| Tertiary Level |  | 41 (11%) |
| **Religion** |  |  |
| Muslim |  | 52 (14%) |
| Christians |  | 311 (85%) |
| Pagans |  | 2 (0.5%) |
| None |  | 2 (0.5%) |
| **Occupation** |  |  |
| Farming |  | 62 (17%) |
| Business |  | 72 (20%) |
| Formal Employment |  | 25 (6.8%) |
| Informal Employment |  | 77 (21%) |
| Pupils / Students |  | 23 (6.3%) |
| Unemployed |  | 108 (29%) |
| **Place of Residence** |  |  |
| Rural |  | 177 (48%) |
| Urban |  | 190 (52%) |

<sup>1</sup>n (%)

**Table 2:** Bivariate Analysis of Factors associated with TB Stigma.

| Characteristic | N | Overall<br>N = 228<sup>1</sup> | No Stigma<br>N = 204<sup>1</sup> | Stigma<br>N = 24<sup>1</sup> | p-value<sup>2</sup> |
| --- | --- | --- | --- | --- | --- |
| **Age** | 228 |  |  |  | 0.75 |
| 0-14 |  | 6 (2.6%) | 6 (2.9%) | 0 (0%) |  |
| 15-24 |  | 37 (16%) | 34 (17%) | 3 (13%) |  |
| 25-34 |  | 64 (28%) | 54 (26%) | 10 (42%) |  |
| 35-44 |  | 59 (26%) | 52 (25%) | 7 (29%) |  |
| 45-54 |  | 28 (12%) | 27 (13%) | 1 (4.2%) |  |
| 55-64 |  | 21 (9.2%) | 19 (9.3%) | 2 (8.3%) |  |
| 65+ |  | 13 (5.7%) | 12 (5.9%) | 1 (4.2%) |  |
| **Sex** | 228 |  |  |  | 0.25 |
| Male |  | 151 (66%) | 138 (68%) | 13 (54%) |  |
| Female |  | 77 (34%) | 66 (32%) | 11 (46%) |  |
| Intersex |  | 0 (0%) | 0 (0%) | 0 (0%) |  |
| **Marital Status** | 228 |  |  |  | 0.65 |
| Single |  | 68 (30%) | 60 (29%) | 8 (33%) |  |
| Married |  | 116 (51%) | 102 (50%) | 14 (58%) |  |
| Divorced |  | 5 (2.2%) | 5 (2.5%) | 0 (0%) |  |
| Widowed |  | 14 (6.1%) | 12 (5.9%) | 2 (8.3%) |  |
| Separated |  | 18 (7.9%) | 18 (8.8%) | 0 (0%) |  |
| Not Applicable |  | 7 (3.1%) | 7 (3.4%) | 0 (0%) |  |
| **Education Level** | 228 |  |  |  | 0.15 |
| No formal education |  | 24 (11%) | 24 (12%) | 0 (0%) |  |
| Primary level |  | 100 (44%) | 90 (44%) | 10 (42%) |  |
| Secondary Level |  | 76 (33%) | 64 (31%) | 12 (50%) |  |
| Tertiary Level |  | 28 (12%) | 26 (13%) | 2 (8.3%) |  |
| **Religion** | 228 |  |  |  | 0.075 |
| Muslim |  | 40 (18%) | 34 (17%) | 6 (25%) |  |
| Christians |  | 186 (82%) | 169 (83%) | 17 (71%) |  |
| Pagans |  | 1 (0.4%) | 1 (0.5%) | 0 (0%) |  |
| None |  | 1 (0.4%) | 0 (0%) | 1 (4.2%) |  |
| **Occupation** | 228 |  |  |  | 0.16 |
| Farming |  | 41 (18%) | 39 (19%) | 2 (8.3%) |  |
| Business |  | 50 (22%) | 45 (22%) | 5 (21%) |  |
| Formal Employment |  | 14 (6.1%) | 11 (5.4%) | 3 (13%) |  |
| Informal Employment |  | 47 (21%) | 38 (19%) | 9 (38%) |  |
| Pupils / Students |  | 17 (7.5%) | 16 (7.8%) | 1 (4.2%) |  |
| Unemployed |  | 59 (26%) | 55 (27%) | 4 (17%) |  |
| **Residence** | 228 |  |  |  | 0.30 |
| Rural |  | 118 (52%) | 108 (53%) | 10 (42%) |  |
| Urban |  | 110 (48%) | 96 (47%) | 14 (58%) |  |
| **Family Size** | 228 | 4.00 (0.00,<br>23.00) | 4.00 (0.00,<br>23.00) | 5.00 (0.00,<br>8.00) | 0.57 |
| **Social Support** | 228 |  |  |  | 0.42 |
| Poor |  | 93 (41%) | 80 (39%) | 13 (54%) |  |
| Moderate |  | 117 (51%) | 107 (52%) | 10 (42%) |  |
| Strong |  | 18 (7.9%) | 17 (8.3%) | 1 (4.2%) |  |
| **Substance use** | 191 |  |  |  | 0.80 |
| Alcohol |  | 26 (14%) | 25 (14%) | 1 (5.9%) |  |
| Miraa (khat) |  | 4 (2.1%) | 4 (2.3%) | 0 (0%) |  |
| Heroin |  | 0 (0%) | 0 (0%) | 0 (0%) |  |
| Tobacco use |  | 3 (1.6%) | 3 (1.7%) | 0 (0%) |  |
| Bhang |  | 1 (0.5%) | 1 (0.6%) | 0 (0%) |  |
| Other (specify) |  | 2 (1.0%) | 2 (1.1%) | 0 (0%) |  |
| None |  | 155 (81%) | 139 (80%) | 16 (94%) |  |
| **Phase** | 222 |  |  |  | 0.089 |
| Intensive phase |  | 84 (38%) | 72 (36%) | 12 (55%) |  |
| Continuation phase |  | 138 (62%) | 128 (64%) | 10 (45%) |  |
| **Patient Type** | 228 |  |  |  | 0.57 |
| New |  | 195 (86%) | 175 (86%) | 20 (83%) |  |
| Relapse |  | 20 (8.8%) | 18 (8.8%) | 2 (8.3%) |  |
| Retreatment |  | 9 (3.9%) | 8 (3.9%) | 1 (4.2%) |  |
| Treatment after Loss to Follow Up |  | 3 (1.3%) | 2 (1.0%) | 1 (4.2%) |  |
| Treatment Failure |  | 1 (0.4%) | 1 (0.5%) | 0 (0%) |  |
| **Mental illness** | 226 | 19 (8.4%) | 18 (8.9%) | 1 (4.2%) | 0.70 |
| **Comorbidities** | 219 |  |  |  | 0.89 |
| HIV |  | 35 (16%) | 32 (16%) | 3 (13%) |  |
| Diabetes |  | 6 (2.7%) | 6 (3.1%) | 0 (0%) |  |
| Cancer |  | 0 (0%) | 0 (0%) | 0 (0%) |  |
| Covid |  | 2 (0.9%) | 2 (1.0%) | 0 (0%) |  |
| Other (specify) |  | 7 (3.2%) | 6 (3.1%) | 1 (4.2%) |  |
| None |  | 169 (77%) | 149 (76%) | 20 (83%) |  |
| **Settings of Stigma** | 178 |  |  |  | <0.001 |
| C1. Hospitals/Clinics |  | 7 (3.9%) | 6 (3.6%) | 1 (8.3%) |  |
| C2. Community/Neighbours |  | 27 (15%) | 20 (12%) | 7 (58%) |  |
| C3. Home/Family |  | 4 (2.2%) | 4 (2.4%) | 0 (0%) |  |
| C4. Workplace |  | 10 (5.6%) | 8 (4.8%) | 2 (17%) |  |
| C5. Prison |  | 13 (7.3%) | 13 (7.8%) | 0 (0%) |  |
| C6. Rescue Centres |  | 1 (0.6%) | 1 (0.6%) | 0 (0%) |  |
| C7. Others |  | 1 (0.6%) | 1 (0.6%) | 0 (0%) |  |
| C8. None |  | 115 (65%) | 113 (68%) | 2 (17%) |  |
| **TB Type** | 225 |  |  |  | 0.34 |
| DSTB |  | 186 (83%) | 168 (84%) | 18 (75%) |  |
| DRTB |  | 5 (2.2%) | 4 (2.0%) | 1 (4.2%) |  |
| Don't Know |  | 34 (15%) | 29 (14%) | 5 (21%) |  |

<sup>1</sup>n (%); Median (Min, Max)
<sup>2</sup>Fisher's exact test; Pearson's Chi-squared test; Wilcoxon rank sum test

##### Informed Consent

All eligible participants were provided with detailed information about the study’ s purpose, procedures, potential risks and benefits, confidentiality measures, and their rights, including the right to decline or withdraw at any time without penalty. Written informed consent was obtained from all participants prior to enrollment. Only those who signed the consent form were included in the study.

## 3. Results

### Participant Demographics

A total of 367 individuals with tuberculosis participated in the study (Table 1), with the largest age group being 25–34 years (30%). Most participants were male (67%) and resided in rural areas (52%), while 48% lived in urban settings. In terms of education, 42% had completed primary school, and 9.8% reported having no formal education. Half of the participants were married, and the majority (85%) identified as Christian. Regarding employment status, 29% were unemployed.

### Tuberculosis-related Stigma Dimensions

The stigma radar was used to assess the level of stigma among various study respondents across five dimensions: self-stigma (patient experience), and secondary stigma from family, community, healthcare settings, and workplaces. As shown in Figure 2, 49% of people with TB (PWTB) reported experiencing self-stigma, with 12% stating it had prevented them from seeking or accessing TB services. Secondary stigma within the family was reported by 52% of participants; however, only 14% indicated it posed a barrier to care.

**Figure 2.**
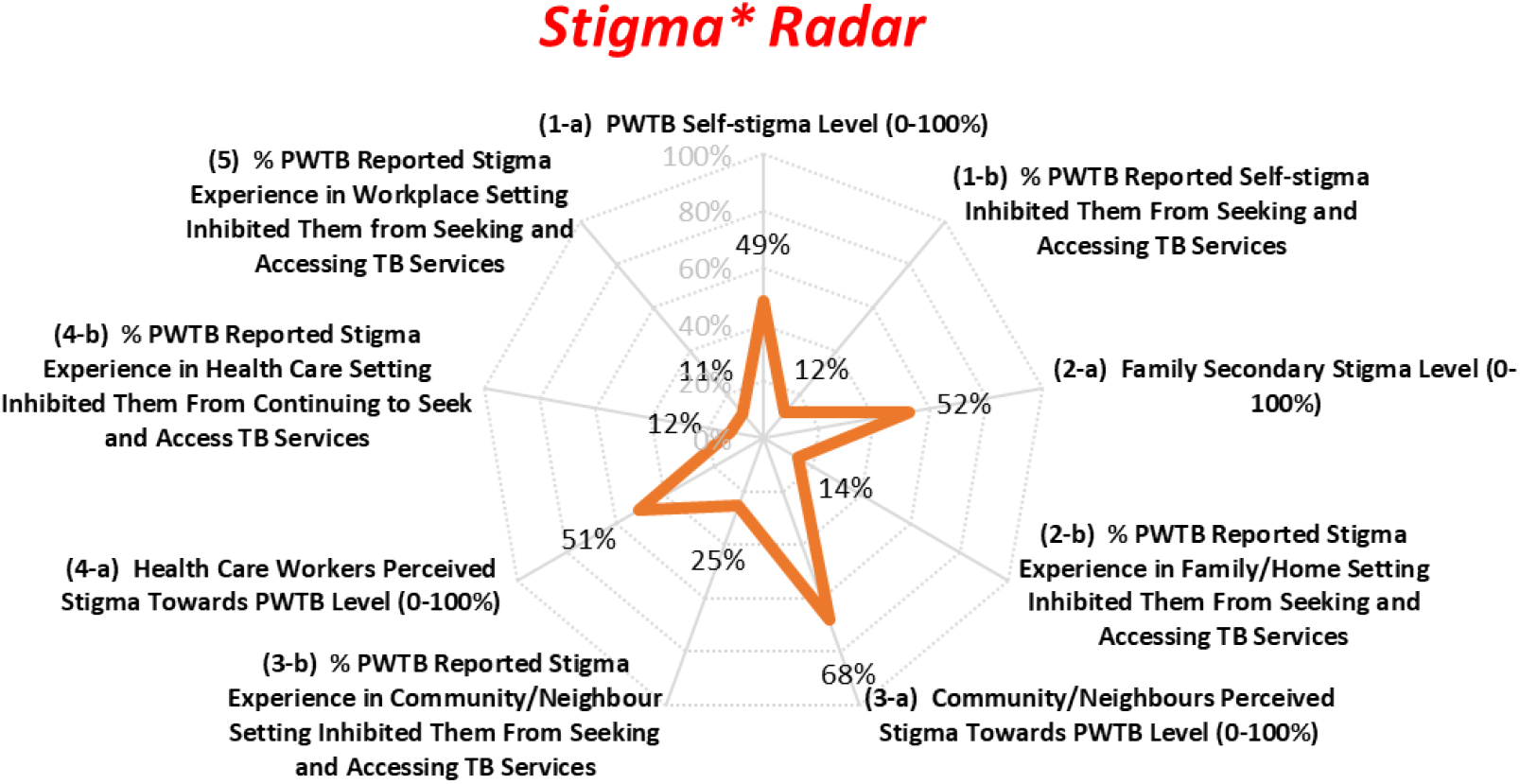
Stigma Radar* adapted from the Stop TB Partnership stigma assessment handbook (26).

Community-level stigma was most prevalent, with 68% of respondents reporting stigma from neighbors or the broader community, and 25% stating that it negatively impacted their access to TB services. Perceived stigma within healthcare settings was reported by 51% of participants, with 12% identifying it as a barrier to care. Workplace-related stigma was noted by 11% of respondents as a deterrent to seeking TB services.

### Factors associated with TB stigma

Table 2 below summarizes the social demographic, and clinical factors associated with TB stigma. Out of 367 participants, 228 individuals with TB shared their experiences regarding stigma. Among them, 24 (11%) reported experiencing TB-related stigma, while 204 (89%) did not. The remaining 139 participants did not provide an opinion and were excluded from the bivariate analysis.

Most demographic and clinical characteristics—including age, sex, marital status, education, occupation, residence, substance use, treatment phase, mental illness, and comorbidities, were not statistically associated with reported stigma (all *p* > 0.05). However, the setting in which stigma was experienced showed a significant association (*p* < 0.001). Among those who reported stigma, 58% identified the community or neighborhood as the primary source, making it the most prevalent setting in which stigma was experienced. Stigma in healthcare settings (8.3%) and workplaces (17%) was less commonly reported, and no respondents reported experiencing stigma at home, in prisons, or in rescue centers.

While not statistically significant, stigma was most frequently reported among individuals aged 25– 44 years (71%), males (54%), and those with secondary-level education (50%). Informal workers and business persons represented the highest proportions of employed individuals reporting stigma. Additionally, those in the intensive phase of TB treatment reported more stigma (55%) than those in the continuation phase (45%), though this difference did not reach statistical significance (*p* = 0.089).

## 4. Discussion

This national cross-sectional study explored the demographic characteristics, levels, and drivers of tuberculosis (TB)-related stigma in Kenya, with a focus on patient experiences across community, family, and healthcare settings. The findings reinforce the multidimensional nature of stigma and offer valuable insights for designing targeted interventions to mitigate stigma and improve treatment outcomes. Among the 367 people with TB (PWTB) surveyed, nearly half reported experiencing some form of stigma, with community-level stigma being the most prevalent. However, only 11% were classified as experiencing significant stigma based on the binary composite score, suggesting that while stigma is widespread in perception, its intensity may vary.

### Demographic characteristics

The demographic characteristics of study participants aligned with national TB surveillance data, with the majority aged 25–44 years (55%) and predominantly male (67%), reflecting global patterns where working-age men bear a higher TB burden (27). Over half of the participants resided in rural areas (52%), and 80% were on drug-sensitive TB treatment, though 14% were unaware of their treatment category, underscoring persistent gaps in patient education.

Although stigma reporting varied by age, sex, education, and residence, none of these were statistically significant predictors. This contrasts with findings from other contexts. In Uganda, higher education was paradoxically linked to greater stigma, possibly due to heightened awareness of TB/HIV associations (18). Conversely, in Colombia, low education and homelessness were associated with increased stigma (28). Informal employment was the most common occupation among those reporting stigma (38%), followed by business owners (21%). Although not statistically significant, this trend is consistent with evidence from Uganda and Colombia, where unemployment and homelessness heightened vulnerability to stigma (18) (28).

### Levels of stigma

Stigma was most pronounced in community settings (68%), followed by family (52%), healthcare settings (51%), and internalized stigma (49%). These findings mirror previous studies from Kenya, South Africa and Uganda, where community level stigma and internalized stigma were the most common forms of stigma reported (12) (29) (30)

These results emphasize stigma as both a psychosocial burden and a critical social determinant of health. The pervasiveness of community-level stigma highlights the urgent need for targeted education campaigns, normalization of TB within communities, and protective strategies for those affected.

### Drivers of Tuberculosis-Related Stigma

Community settings emerged as the leading source of stigma, with 58% of participants reporting negative experiences. This reinforces findings from South Africa, where gossip, visible symptoms, perceived links to HIV, and fear of infection are major drivers of exclusion (29) (31)(32).

Stigma was elevated during the intensive treatment phase (55%), consistent with South Africa studies linking visible symptoms during early treatment to concealment and withdrawal (33). Patients with drug-resistant TB also reported higher levels of stigma, reflecting the prolonged treatment duration and perceptions of incurability (34) (35). Religious context also influenced stigma, with both Christians and Muslims reporting experiences stigma. These findings align with those that indicated that the role of religion in stigma appears to vary by setting (36).

Social support emerged as an important protective factor: participants with poor support reported higher stigma (54%). This aligns with studies from China, Tanzania and Kenya which demonstrate that strong social support reduces stigma experiences (12) (37) (38). While education was not significantly associated with stigma in this study, evidence from sub-Saharan Africa suggests that higher education and wealth are protective, reducing the likelihood of concealing TB status and lowering self-stigma (36) (20).

## 5. Conclusion

This national survey highlights the widespread and multifaceted nature of TB-related stigma in Kenya, with community settings emerging as the most significant source of stigma, particularly during early treatment phases. Stigma was shaped by demographic and social factors, including occupation, religion, social support, and treatment type, often resulting in delayed disclosure and care-seeking. These findings underscore the urgent need for targeted, community-driven interventions that address misconceptions, foster supportive environments, and integrate psychosocial support across the TB care cascade to improve patient outcomes and advance TB elimination goals.

## Data Availability

The data will be made readily available with any analytic scripts as may be required

## Acknowledgments

The successful completion of this study was made possible through the collective efforts and dedication of numerous individuals and organizations. Gratitude is extended to people with TB for sharing their experiences, healthcare workers for their support in planning interview appointments with patients. Researchers, data analysts, and NTP officers ensured accurate findings, while government agencies, NGOs, and international partners provided essential support. Community leaders and advocates are also recognized for their commitment to reducing TB stigma. Their combined efforts help create a stigma-free environment for TB-affected individuals.

